# The general neurocognitive decline in patients with methamphetamine (MA) use and transient MA-induced psychosis is primarily determined by oxidative and AGE-RAGE stress

**DOI:** 10.1101/2024.04.02.24305184

**Authors:** Michael Maes, Mazin Fadhil Altufaili, Amer Fadhil Alhaideri, Shatha Rouf Moustafa, Kristina Stoyanova, Mengqi Niu, Bo Zhou, Jing Li, Hussein Kadhem Al-Hakeim

**Affiliations:** Sichuan Provincial Center for Mental Health, Sichuan Provincial People’s Hospital, School of Medicine, University of Electronic Science and Technology of China, Chengdu 610072, China; Key Laboratory of Psychosomatic Medicine, Chinese Academy of Medical Sciences, Chengdu, 610072, China; Department of Psychiatry, Faculty of Medicine, Chulalongkorn University, Bangkok, Thailand; Cognitive Impairment and Dementia Research Unit, Faculty of Medicine, Chulalongkorn University, Bangkok, Thailand; Cognitive Fitness and Biopsychological Technology Research Unit, Faculty of Medicine Chulalongkorn University, Bangkok, 10330, Thailand, Bangkok, 10330, Thailand; Department of Psychiatry, Medical University of Plovdiv, Plovdiv, Bulgaria; Research Institute, Medical University of Plovdiv, Plovdiv, Bulgaria; Kyung Hee University, 26 Kyungheedae-ro, Dongdaemun-gu, Seoul 02447, Korea; Research and Innovation Program for the Development of MU - PLOVDIV–(SRIPD-MUP)“, Creation of a network of research higher schools, National plan for recovery and sustainability, European Union – NextGenerationEU; Department of Chemistry, College of Science, University of Kufa, Iraq.; Senior Psychiatrist at the College of Medicine, University of Kerbala, Iraq.; Clinical Analysis Department, College of Pharmacy, Hawler Medical University, Erbil, Iraq.; Department of Chemistry, College of Science, University of Kufa, Kufa 54002, Iraq.

**Keywords:** pathways, schizophrenia, oxidative and nitrosative stress, antioxidants, biomarkers

## Abstract

**Background:** Chronic methamphetamine (MA) usage is linked to oxidative stress (OS), AGE-RAGE stress, changes in magnesium, calcium, and copper, increased psychotic symptoms and neurocognitive deficits. Nevertheless, it is still unclear whether the latter impairments are mediated by these biological pathways.

**Aims:** The purpose of this study was to investigate the relationships between neurocognition, the aforementioned biomarkers, and psychotic symptoms.

**Methods:** We recruited 67 participants, namely 40 patients diagnosed with MA-substance use and 27 healthy controls, and assessed the Brief Assessment of Cognition in Schizophrenia (BACS), symptoms of psychosis, excitation, and formal thought disorders, OS toxicity (computed as the sum of myeloperoxidase (MPO), oxidized high-density lipoprotein (HDL), oxidized low-DL, and malondialdehyde), antioxidant defenses (catalase, glutathione peroxidase, total antioxidant capacity, zinc, and HDL), increased advanced glycation end products (AGEs), and soluble AGE receptors.

**Results:** We were able to extract one validated latent vector from the Mini Mental State Examination score and the BACS tests results (including executive functions, verbal fluency, attention), labeled general cognitive decline (G-CoDe). We found that 76.1% of the variance in the G-CoDe was explained by increased OS toxicity, lowered antioxidant defenses, number of psychotic episodes, and MA dose. In patients with MA use, MPO was significantly associated with the G-CoDe.

**Conclusions:** The use of MA induces mild cognitive impairments through MA-induced activation of detrimental outcome pathways, including oxidative and AGE-RAGE stress, and suppression of protective outcome pathways (antioxidants). Increased OS, MPO, and AGE-RAGE stress are new drug targets to prevent neurocognitive deficits and psychosis due to MA use.

## Introduction

Derived from amphetamine, methamphetamine (MA) is a potent psychostimulant that is the second most abused substance after cannabis [1-5]. MA is often used to combat fatigue, boost pleasure, enhance social interactions, increase libido, improve productivity, and suppress appetite for weight loss purposes [6,7]. MA abuse is a major global health concern because of its easy accessibility, cheap price, addictive nature, and rising incidents of overdose-related complications and deaths [8]. Moreover, when people use MA, it can result in a range of neuropsychiatric symptoms and behaviors, including affective and somatic manifestations, psychosis, and neurocognitive deficits [9-15].

Typical symptoms of psychosis caused by MA include hallucinations, having paranoid beliefs, and thinking that unrelated events are connected, as shown in various research [8,9,14,16,17]. Research indicates that 40% of MA users may exhibit psychotic symptoms and cognitive symptoms resembling those observed in schizophrenia, indicating that MA-induced psychosis (MIP) could serve as a model for schizophrenia. Based on a meta-analysis, the occurrence of MIP is 36.5%, with a lifetime occurrence of 42.7% [18]. Factors that increase the risk of MIP include MA dependence, frequent and heavy use of methamphetamine, MA dosage levels [19]. MIP is typically a transient condition that lasts a few days, although 5-15% of users may suffer from more persistent symptoms [20].

It is common for individuals with MA abuse and dependence to exhibit deficits in different neurocognitive functions and psychomotor skills [15]. These impairments may be linked to continuous drug use and the level of dependence [15]. Dysfunctional cognitive domains include set-shifting performance, memory and recall, and response inhibition. Based on the available data, acute MA use has been shown to improve cognitive performance in attention and visuospatial perception [21]. Upon examining the effects of chronic MA abuse on cognitive performance, researchers have found notable distinctions between individuals who use MA and those in the control group. However, these cognitive impairments typically fall within normal limits [21]. The authors suggest that there may have been an overinterpretation of positive findings. In 2018, a literature review by Potvin et al. [22] revealed moderate impairments in executive functions, attention, memory, verbal fluency, and learning, working memory. However, visuo-spatial abilities and visual learning were relatively unaffected.

We have recently published findings that show a connection between the severity of MA-use, MA-dependence, and MIP with various biomarkers such as advanced glycation end products (AGEs), soluble AGE receptors (sRAGE), the atherogenic index of plasma (AIP), oxidative and nitrosative stress (O&NS) indicators, aberrations in the cytokine-chemokine-growth factor network, and lowered levels of zinc, calcium and magnesium [8,16,17]. One important finding was that AGE and sRAGE levels, O&NS index, magnesium levels, and copper levels together explained 54.8% of the variability in psychotic symptoms and were found to mediate the impact of increasing MA dose on MIP symptoms [17]. We have detailed in previous studies [16,17] how various factors such as oxidative stress, antioxidant defenses, AGE-RAGE stress, and magnesium and copper levels can contribute to neurotoxicity and increase the risk of MIP. A consistent pattern has been identified in the most severe subtype of schizophrenia (“major neurocognitive psychosis”), where heightened neurotoxicity of various biomarkers (including O&NS) is intricately linked to the intensity of psychosis and neurocognitive impairments such as semantic and episodic memory, executive functions, and attention [23,24,25].

However, it is still unclear - in patients with MA use - whether O&NS, AGE-RAGE stress, and alterations in magnesium and copper levels are linked to cognitive impairments, and if these mechanisms play a role in the impact of MA dosage and dependence on neurocognitive deficits. Furthermore, it has been unclear whether a single overarching factor can be derived from various cognitive test outcomes, including working memory, verbal episodic memory, semantic fluency, letter fluency, attention, and executive functions, as demonstrated in schizophrenia research [25]. This general factor was labeled “general cognitive decline” (G-CoDe). Hence, this study aimed to investigate if a single latent construct, namely a G-CoDe, could be identified for different neurocognitive impairments in individuals with MA use. Additionally, the study examined whether the biomarkers mentioned could predict G-CoDe and mediate the impact of MA dose or dependence on cognitive deficits.

### Participants

The 67 participants in this investigation were comprised of 40 patients diagnosed with MA-substance use disorder (SUD) and 27 healthy controls. The study was conducted at the Psychiatry Unit of Al-Hussein Medical City in the Kerbala Governorate, Iraq, spanning from April to August 2022. MA-SUD was identified as moderate to severe MA substance use disorder (SUD) in accordance with the Diagnostic and Statistical Manual of Mental Disorders, 5th edition (DSM-5) [26] and MA SUD (F15.20) in the International Classification of Diseases. Due to the prevailing religious atmosphere in Karbala, our recruitment efforts were limited to male addicts. At least three months of MA abuse was documented in every MA-SUD patient, and a urine MA test was positive for the drug. We included only patients without MIP and those with transient MIP not lasting more than 4 days after admission into hospital. The healthy controls comprised individuals who were known to either the personnel or the patients.

The same study sample was used in our previous studies [16,17]. However, in order to examine neurocognitive deficits, we excluded in the current study illiterate participants. Exclusion criteria also encompassed patients and controls diagnosed with (auto)immune disorders such as inflammatory bowel disease, rheumatoid arthritis, chronic obstructive pulmonary disease, diabetes mellitus, or psoriasis. Patients and controls with a history of immunomodulatory drug treatment (e.g., glucocorticoids, immunosuppressive drugs), as well as those with neurodegenerative or neuroinflammatory disorders (e.g., stroke, multiple sclerosis, Alzheimer’s disease, Parkinson’s disease, or multiple sclerosis), were excluded from the study. In both patients and controls, there was no evidence of dependence on any other substances among the patients, such as cocaine, opium, or opioids. Furthermore, it is important to note that none of the control subjects exhibited any known familial history of schizophrenia or psychosis, nor had any of them been diagnosed with an axis I DSM-IV-TR disorder (with the exception of nicotine dependence).

The investigation was conducted in accordance with global and Iraqi ethical and privacy standards. All participants and first-degree relatives of participants with MA SUD (whose legal representatives are the mother, father, brother, spouse, or son) provided written informed consent prior to their involvement in this study. Following the International Guideline for Human Research Protection of the Declaration of Helsinki, the study was approved by the institutional review board (IRB) of the College of Science, University of Kufa, Iraq (89/2022), Karbala Health Directorate-Training and Human Development Center (Document No.18378/2021).

## Clinical evaluations

Clinician and demographic information were gathered on the same day that blood samples were drawn and the day following hospital admission. In pursuit of this objective, a senior psychiatrist specializing in addiction administered rating scales and conducted a semi-structured interview to evaluate the severity of MA dependence, use, and psychosis. Age at onset, duration of MA dependence, daily dosage (grammes), route of administration (ordinal variable with no=0, inhaled or snorted=2, and injected=3) were also recorded. The patients were categorized into two categories based on the quantity of MA used: the high-MA SUD group (consisting of 31 patients who consumed 2.435±0.616 g of MA daily) and the low-MA SUD group (comprising 29 patients who consumed 0.948±0.155 g of MA daily). Additionally, we documented both chronic cannabis and alcohol use, in addition to dependence on both substances.

The number of prior episodes of psychosis and the duration of hospitalization subsequent to admission for acute intoxication or psychosis were recorded. The severity of MA dependence was assessed using the Severity of the Dependence Scale (SDS) [27], which consisted of the following five items: a) Have you ever considered that your MA use was unmanageable? b) Does the thought of going without MA cause you significant anxiety or concern? c) Do you harbor concerns regarding your MA use? d) Do you wish you could discontinue MA? and e) To what extent would you find it challenging to cease or go without MA?

We evaluated the Positive and Negative Syndrome Scale (PANSS) [28] and the Brief Psychiatric Rating Scale (BPRS) [29] on the same day as blood sampling and the clinical interviews. As previously described [16,17], we calculated various z unit-weighted composite scores to assess the severity of psychosis using items from the BPRS and PANNS: (a) psychosis was determined by summing the z transformations of delusions (PANSP1) + hallucinatory behavior (PANSS P3) + suspiciousness (PANNS P6); and (b) excitement was determined by summing the z transformations of grandiosity (BPRS), excitement (BPRS), suspiciousness (BPRS), and grandiosity (PANNS P4). We performed cluster analysis on increased psychosis and excitation composite scores to identify a cohort of patients with increased psychosis symptoms (refer to the section below).

The neurocognitive tests conducted by a clinical psychologist who was unaware of the diagnosis and clinical ratings utilized the Brief Assessment of Cognition in Schizophrenia (BACS) [30]. These examinations were conducted in the absence of any clinical manifestations of psychosis. The interval between admission and neurocognitive testing performance was 0–6 days (mean=1.52 days; q25=0, q75=3 days). The BACS screening consists of the following assessments: working memory (Digit Sequencing Task), verbal episodic memory (List Learning test), semantic fluency (Controlled oral Word Association test or COWAT), and letter fluency (Controlled Word Association); attention (Symbol Coding); and executive functions (Tower of London). Utilization of tobacco (TUD) was diagnosed in accordance with DSM-5 criteria. The body mass index (BMI) was calculated utilizing the subsequent formula: body weight (kg) divided by length (m2).

## Methods

The urine MA test was conducted upon admission with the Multi Drug 12 Drugs Rapid Test Panel (Urine) kit provided by Citest Diagnostics Inc. (Vancouver, Canada). On the morning following admission, fasting venous blood samples were collected from all participants upon their arrival at the hospital. The blood was left to coagulate for 10 minutes at room temperature and then centrifuged at 3000 rpm for 10 minutes. The serum was carefully separated and transferred to Eppendorf tubes for storage at -80 °C until analysis. High-density lipoprotein cholesterol (HDL) levels were assessed with a kit provided by Spinreact®, located in Gerona, Spain. Serum zinc levels were analyzed spectrophotometrically using a ready-to-use kit from Agappe Diagnostics® based in Cham, Switzerland. The concentration of copper was analyzed using a spectrophotometric kit from LTA Co (Milan, Italy). The serum calcium and magnesium concentrations were analyzed using spectrophotometric kits from Biolabo® (Maizy, France). Levels of glutathione peroxidase (Gpx), myeloperoxidase (MPO), malondialdehyde (MDA), oxidized HDL (OxHDL), oxidized LDL (OxLDL), total antioxidant capacity (TAC), catalase, AGE, and sRAGE were determined using commercial ELISA kits provided by Nanjing Pars Biochem Co., Ltd. (Nanjing, China). All kits utilized a sandwich technique and demonstrated an inter-assay CV of less than 10%. As previously discussed, we utilized a range of composite scores: a) a composite score representing the AGE-RAGE axis stress was derived from z AGE + z sRAGE, referred to as AGE-RAGE axis (ARA) stress; b) OSTOX was determined by z MPO + z oxHDL + z oxLDL + z MDA; c) ANTIOX was calculated using z catalase + z Gpx + z TAC + z zinc + z HDL; and d) The OSTOX/ANTIOX ratio was computed as z OSTOX – z ANTIOX.

## Analysis of statistics

The Pearson’s product-moment or Spearman’s rank order correlation coefficient was used to analyze the relationship between two scale variables. Utilizing analysis of variance (ANOVA) to compare groups in continuous variables and contingency tables (χ2-test) to explore relationships among nominal variables. We employed a multivariate general linear model (GLM) to analyze the connections between study groups (healthy control and patient groups) and the psychiatric rating scale scores composites and biomarkers, while accounting for confounding variables such as age, sex, smoking, and education. We computed the estimated marginal mean values (SE) and used protected least significant difference (LSD) tests to conduct pairwise comparisons among the group means, with the omnibus test being significant. We also utilized false discovery rate (FDR) p-correction for the multiple comparisons [31].

Furthermore, multiple regression analysis has been used to investigate if the biomarkers can accurately predict the results of the neurocognitive tests. We utilized both a manual method and a stepwise automated approach, with specific p-values for entry and removal from the model (p=0.05 and p=0.06, respectively). We calculated standardized beta coefficients along with t statistics and exact p-values for each predictor variable. Additionally, we calculated model statistics (F, df, and p values) and total variance explained (R^2^) as the effect size. In addition, the variance inflation factor (VIF) and tolerance were utilized to assess collinearity and multicollinearity concerns. We conducted tests for heteroskedasticity using the White and modified Breusch-Pagan homoscedasticity tests and, when needed, applied univariate GLM analysis to estimate parameters. Binary logistic regression analysis was conducted (manual and automatic step up) to delineate the most important predictors. We computed Wald statistics with exact p-value and Odd’ratios with 95% confidence intervals. The adequacy of the regression model was checked using the accuracy and Nagelkerke values. The results of the regression analyses were consistently bootstrapped with 5,000 bootstrap samples, and the latter were disclosed if the results were not in agreement. Two-tailed tests were utilized to assess significance, with a significance level set at p=0.05 in SPSS version 28 (windows) for all statistical analyses.

A cluster analysis was conducted using a two-step cluster approach, considering both categorical and continuous variables. The cluster solution’s adequacy was assessed by examining the silhouette measure of cohesion and separation. It was considered acceptable if it surpassed the threshold of 0.3. Principal Components Analysis (PCA) was utilized to extract latent constructs from a set of variables (e.g., all 8 cognitive tests results). The Kaiser-Meier-Olkin (KMO) sample adequacy measure was utilized to evaluate factorability, which is considered sufficient when it exceeds 0.7. In addition, if all loadings on the first factor exceeded 0.66, the variance explained by the first factor was over 50%, and Cronbach’s alpha calculated on the variables was above 0.7, the first principal component was considered a valid latent construct supporting the variables.

An analysis using Partial Least Squares (PLS) was performed to examine the causal relationships between the use/dose of MA, biomarkers, and cognitive test results. A single factor was extracted from the cognitive test results and used as the output variable. Complete PLS analysis is conducted only under specific conditions: The results of the Confirmatory Tetrad Analysis (CTA) should demonstrate that the factor constructed is accurately described as a reflective model. The PLSpredict analysis indicates that the model’s predictive performance is efficient. The factor extracted exhibits adequate construct and convergence validity, as indicated by composite reliability > 0.8, Cronbach’s alpha > 0.7, and average variance extracted (AVE) > 0.5. Additionally, all loadings on the latent vectors should be > 0.66 at p < 0.001. Lastly, the model demonstrates adequate fit, with standardized root squared residual (SRMR) values < 0.08. By utilizing 5,000 bootstraps, we have calculated path coefficients, including their corresponding p-values. Additionally, we have determined the specific and total indirect effects, as well as the total effects. Based on a priori power analysis, it has been determined that the estimated sample size required for the primary analysis (Partial Least Squares (PLS) analysis) is 58 participants. The power level is set at 0.8, the significance level (alpha) is 0.05, and the effect size is 0.25 (around 20% of the variance explained). Additionally, the analysis will involve a maximum of 5 predictors.

## Results

### Sociodemographic and clinical data

In order to define a cluster with increased psychosis scores we performed two step cluster analysis with the diagnosis MA-use versus controls as categorical variable and psychosis and excitation as continuous variables. A three-cluster solution showed a silhouette measure of cohesion and separation of 0.76, indicating a high cluster quality. Three clusters were formed, a normal control cluster and two clusters of patients with MA abuse, one with severe psychosis + excitation scores (labeled MA-PES) and one with lower scores (labeled MA).

**Table 1** shows the sociodemographic and clinical data of patients with MA and MA-PES and healthy controls. There were no significant differences in age, education, and BMI between the three study groups. The unemployment ratio was higher in MA-PES. Duration of disease and the SDS score was higher in MA-PES patients than in those without MA-PES. There was a trend towards increased MA dose in MA-PES. The prevalence of alcohol use and TUD were higher in patients than in controls, especially in MA-PES patients. All neurocognitive test scores were significantly lower in patients than in controls, while COWAT, symbol coding was lower in MA-PES patients than in patients without.

**Table 1.** Sociodemographic and clinical data of patients with MA abuse and MA-induced psychosis and excitation symptoms (PES) and healthy controls. All data are shown as mean ±SD. F/Χ^2^: results of analysis of variance and chi-square test (contingency analysis, respectively). a,b,c: comparison among group means. G-CoDe: general cognitive decline computed based on the Mini Mental State Examination (MMSE) and cognitive tests as shown in Table 1, FTD+G-CoDE: factor based on formal thought disorders and the G-CoDe tests; FPE+G-CoDe: factor based on formal thought disorders, psychosis, excitation and the G-CoDe tests (see Table 2); OSTOX: oxidative stress toxicity; ANTIOX: antioxidant defenses; sRAGE: soluble receptors for advanced glycation end products (AGEs).

| Variables | HC a | MA b | MA-PES c | F/X<sup>2</sup> | df | p |
| --- | --- | --- | --- | --- | --- | --- |
| Age (years) | 26.5 ±5.2 | 26.1 ±4.9 | 27.4 5.5 | 0.33 | 2/64 | 0.719 |
| Education (years) | 11.4 ±3.5 | 13.2 ±2.9 | 13.8 4.1 | 2.91 | 2/64 | 0.061 |
| Employment (No/Yes) | 6/21 | 8/14 | 13/5 | 11.43 | 2 | 0.003 |
| BMI | 23.32 2.96 | 25.12 ±3.87 | 25.27 3.18 | 0.02 | 2/64 | 0.977 |
| Duration of disease | - | 17.9 ±15.9 c | 32.1 17.2 a | 7.30 | 1/38 | 0.010 |
| MA dose (mg/day) | - | 1.47 ±0.79 c | 1.97 0.87 a | 3.55 | 1/38 | 0.067 |
| MA route 0/1/2/3 | 27/0/0/0/ | 6/9/4/3 | 5/4/9/0 | 45.57 | 6 | <0.001 |
| SDS score | - | 9.41 1.05 | 10.5 | 7.24 | 1/38 | 0.011 |
| Alcohol use (No/Yes) | 27/0 | 15/7 | 3/15 | 34.02 | 2 | <0.001 |
| TUD | 14/13 | 4/18 | 0/18 | 16.04 | 2 | <0.001 |
| MMSE | 29.0 ±1.4 b,c | 25.5 ±2.3 a | 24.7 1.9 a | 35.13 | 2/64 | <0.001 |
| List learning | 61.1 ±5.9 b,c | 51.9 ±4.6 a | 50.3 5.5 a | 27.55 | 2/64 | <0.001 |
| Digit sequencing task | 21.5 ±3.1 b,c | 14.1 ±3.6 a | 14.1 4.0 a | 35.93 | 2/64 | <0.001 |
| Token motor task | 99.4 ±2.8 b,c | 72.1 ±17.3 a | 59.8 16.6 a | 54.17 | 2/64 | <0.001 |
| Category instances | 72.9 ±5.6 b,c | 45.4 ±13.1 a | 41.9 9.2 a | 75.38 | 2/64 | <0.001 |
| COWAT | 35.2 ±9.4 b,c | 23.7 ±6.8 a,c | 17.6 8.4 a,b | 26.11 | 2/64 | <0.001 |
| Symbol coding | 83.9 ±5.5 b,c | 51.1 ±14.0 a,c | 44.3 17.6 a,b | 66.50 | 2/64 | <0.001 |
| Tower of London | 20.1 ±1.5 b,c | 13.7 ±3.8 a | 11.9 5.3 a | 32.34 | 2/64 | <0.001 |
| G-CoDe | 1.099 ±0.306 b,c | -0.568 ±0.426 a,c | -0.955 0.461 a,b | 181.64 | 2/64 | <0.001 |
| Formal thought Disorders (FTD) | -1.138 b,c | 0.310 ±0.740 a,c | 1.080 0.442 a,b | 124.74 | 2/64 | <0.001 |
| FTD+G-CoDe | 1.113 ±0.266 b,c | -0.550 ±0.396 a,c | -0.997 0.414 a,b | 230.52 | 2/64 | <0.001 |
| Psychosis (z score) | -1.170 b,c | 0.349 ±0.717 a,c | 1.126 0.330 a,b | 157.34 | 2/64 | <0.001 |
| Excitation (z score) | -1.038 b,c | -0.150 ±0.536 a,c | 1.244 0.359 a,b | 218.80 | 2/64 | <0.001 |
| FPE-G-CoDe | 1.122 ±0.209 b,c | -0.483 ±0.349 a,c | -1.094 0.308 a,b | 365.43 | 2/64 | <0.001 |

### Results of principal component analysis

**Table 2** shows the results of PC analyses and construction of three different PCs. First, we were able to extract one PC from the 8 cognitive BACS test scores. This PC showed adequate factorability and the first PC explained 64.94% of the data, and all loadings were > 0.7. As such, we constructed a validated PC, named the G-CoDe. Table 1 shows that the G-CoDe factor score was significantly different between the three groups and decreased from controls to patients with MA use to patients with psychotic symptoms. The same table shows that we were able to add FTD to the same PC and to extract one validated PC from the 8 cognitive test scores and FTD. This PC is labeled FTD-GCoDe (labelled: PEF-GCoDe), Finally, we were able to extract one validated PC from the 8 cognitive scores, FTD, psychosis and excitation. Table 1 shows that FTD, psychosis and excitation as well as their combined composites with the G-CoDe were significantly different between the three groups and increased from controls to MA patients to those with MA-PES.

**Table 2.** Results of principal component (PC) analysis and construction of different latent constructs based on cognitive tests, and symptoms of formal thought disorders, psychosis, and excitation. KMO: The Kaiser-Meyer-Olkin Test; VE: Variance explained; Χ^2^: Bartlett’s test of sphericity. MMSE: Mini Mental State Examination; COWAT: Controlled oral Word Association test. G-CoDe: general cognitive decline computed based on the MMSE and cognitive tests; FTD+G-CoDe: factor based on formal thought disorders and the G-CoDe tests; FPE+G-CoDe: factor based on formal thought disorders, psychosis, excitation, and the G-CoDe tests.

| **G-CoDe** |  | **FTD-GCoDe** |  | **FPE-GCoDe** |  |
| --- | --- | --- | --- | --- | --- |
| **Variables** | **Loading** | **Variables** | **Loading** | **Variables** | **Loading** |
| MMSE | 0.791 | MMSE | .790 | MMSE | .785 |
| List learning | 0.760 | List learning | .758 | List learning | .741 |
| Digit sequencing task | 0.785 | Digit sequencing task | .775 | Digit sequencing task | .753 |
| Token motor task | 0.739 | Token motor task | .754 | Token motor task | .775 |
| Category instances | 0.886 | Category instances | .893 | Category instances | .880 |
| COWAT | 0.766 | COWAT | .754 | COWAT | .743 |
| Symbol coding | 0.887 | Symbol coding | .881 | Symbol coding | .868 |
| Tower of London | 0.819 | Tower of London | .805 | Tower of London | .782 |
|  |  | Formal thought disorders | -.870 | Formal thought disorders | -.901 |
|  |  |  |  | Psychosis | -.902 |
|  |  |  |  | Excitation | -.836 |
| KMO = 0.919 |  | KMO = 0.932 |  | KMO = 0.921 |  |
| $X^2 = 336.885$ (df=28), $p < 0.001^{***}$ | | $X^2 = 417.050$ (df=36), $p < 0.001^{***}$ | | $X^2 = 622.585$ (df=55), $p < 0.001^{***}$ | |
| VE = 64.94% |  | VE = 65.74% |  | VE = 66.83% |  |

KMO: The Kaiser-Meyer-Olkin Test; VE: Variance explained; $X^2$ : Bartlett's test of sphericity.

### Intercorrelation matrix between the constructed PCs and the biomarkers

**Table 3** shows the intercorrelation (Pearson’s product moment correlations) between the three constructed PCs and the biomarkers. The three PCs were significantly correlated with all biomarkers, except oxidized HDL and Gpx. The highest correlation coefficients were observed for magnesium, copper, AIP, OSTOX, ANTIOX, and sRAGEs. The highest correlation coefficient was established between the three PCs and the OSTOX/ANTIOX ratio.

**Table 3.** Intercorrelation matrix between neurocognitive test results and biomarkers in subjects with methamphetamine use. All correlations at p<0.01, except °: not significant and * p<0.05 (n=67) G-CoDe: general cognitive decline computed based on the MMSE and cognitive tests; FTD+G-CoDe: factor based on formal thought disorders and the G-CoDe tests; FPE+G-CoDe: factor based on formal thought disorders, psychosis, excitation, and the G-CoDe tests.

| Variables | G-CoDe | FTD-GCoDe | FPE-GCoDe |
| --- | --- | --- | --- |
| Magnesium | 0.587 | 0.595 | 0.609 |
| Calcium | 0.294 * | 0.298 * | 0.301 * |
| Copper | -0.518 | -0.532 | -0.550 |
| Zinc | 0.353 | 0.357 | 0.373 |
| High-density lipoprotein cholesterol (HDL) | 0.372 | 0.369 | 0.366 |
| Oxidized HDL | -0.199 ° | -0.213 ° | -0.210 ° |
| Oxidized Low density lipoprotein cholesterol | -0.450 | -0.440 | -0.417 |
| Malondialdehyde | -0.469 | -0.454 | -0.431 |
| Myeloperoxidase | -0.292 * | -0.292 * | -0.296 * |
| Nitric oxide metabolites | 0.113 | 0.111 | 0.104 |
| Oxidative toxicity (OSTOX) | -0.631 | -0.627 | -0.607 |
| Catalase | 0.279 * | 0.271 * | 0.245 * |
| Glutathione peroxidase | 0.087 ° | 0.102 ° | 0.099 ° |
| Total antioxidant capacity | 0.417 | 0.430 | 0.430 |
| Antioxidant defenses (ANTIOX) | 0.521 | 0.528 | 0.522 |
| OSTOX/ANTIOX | -0.728 | -0.730 | -0.714 |
| Advanced glycation end products (AGEs) | -0.312 * | -0.313 * | -0.297 * |
| Soluble receptors AGEs (sRAGEs) | -0.529 | -0.546 | -0.555 |

### Intercorrelation matrix between clinical cognitive and dependence features

**Table 4** shows the intercorrelations (Spearman’s rank order correlation coefficients) between clinical variables (psychosis, excitation, FTD), G-CoDe and MA use features. These analyses were performed in patients with MA abuse. This table shows that the number of prior psychotic episodes was correlated with the psychosis score. There were no significant correlations between days hospitalized and any of the variables. Alcohol use and the SDS score were significantly correlated with psychosis and excitation scores, but not with FTD ot G-CoDe. As such, none of the dependency variables was associated with the G-CoDe. The MA dose was significantly correlated with psychosis and FTD.

**Table 4.** Intercorrelation matrix between clinical and neurocognitive scores and methamphetamine (MA) use features. All correlations at p<0.01, except °: non-significant (n=40)

| Variables | Psychosis | Excitation | FTD | G-CoDe |
| --- | --- | --- | --- | --- |
| Number of MA-psychoses | **0.439** | 0.152 ° | 0.254 ° | 0.034 ° |
| Days hospitalized | 0.177 ° | -0.173 ° | 0.215 ° | -0.065 ° |
| Alcohol use | **0.523** | **0.409** | 0.276 ° | -0.244 ° |
| MA dose | **0.408** | 0.228 ° | **0.511** | 0.134 ° |
| SDS score | 0.188 ° | **0.404** | 0.038 ° | 0.092 ° |

### Results of multiple regression analysis

**Tables 5-6** show the results of multiple regression analyses with the three PC scores as dependent variables and the biomarkers and demographic data as explanatory variables. Table 5 shows the results obtained on all participants (n=67), and Table 6 shows results obtained in MA patients. Table 5, regression 1a shows that 64.7% of the variance in the G-CoDe was explained by the OSTOX/ANTIOX ratio, sRAGE (both inversely) and magnesium (positively). Table 6 shows that in the selected group of MA patients, MPO was inversely associated with the G-CoDe score, explaining 14.6% of the variance. The results of multiple regression analyses performed om the FTD-GCoDe and PEF-GCoDe scores showed that the same predictors appeared in the analyses. We have also rerun the analysis shown in regression 1a but now with number of episodes included. We found that 70.2% of the variance in the G-CoDe was explained by increased OSTOX/ANTIOX, sRAGE, and number of psychotic episodes. **Figure 1** shows the partial regression of the GCoDe score on the OSTOX/ANTIOX ratio (after controlling for number of episodes and sRAGE).

**Figure 1.**
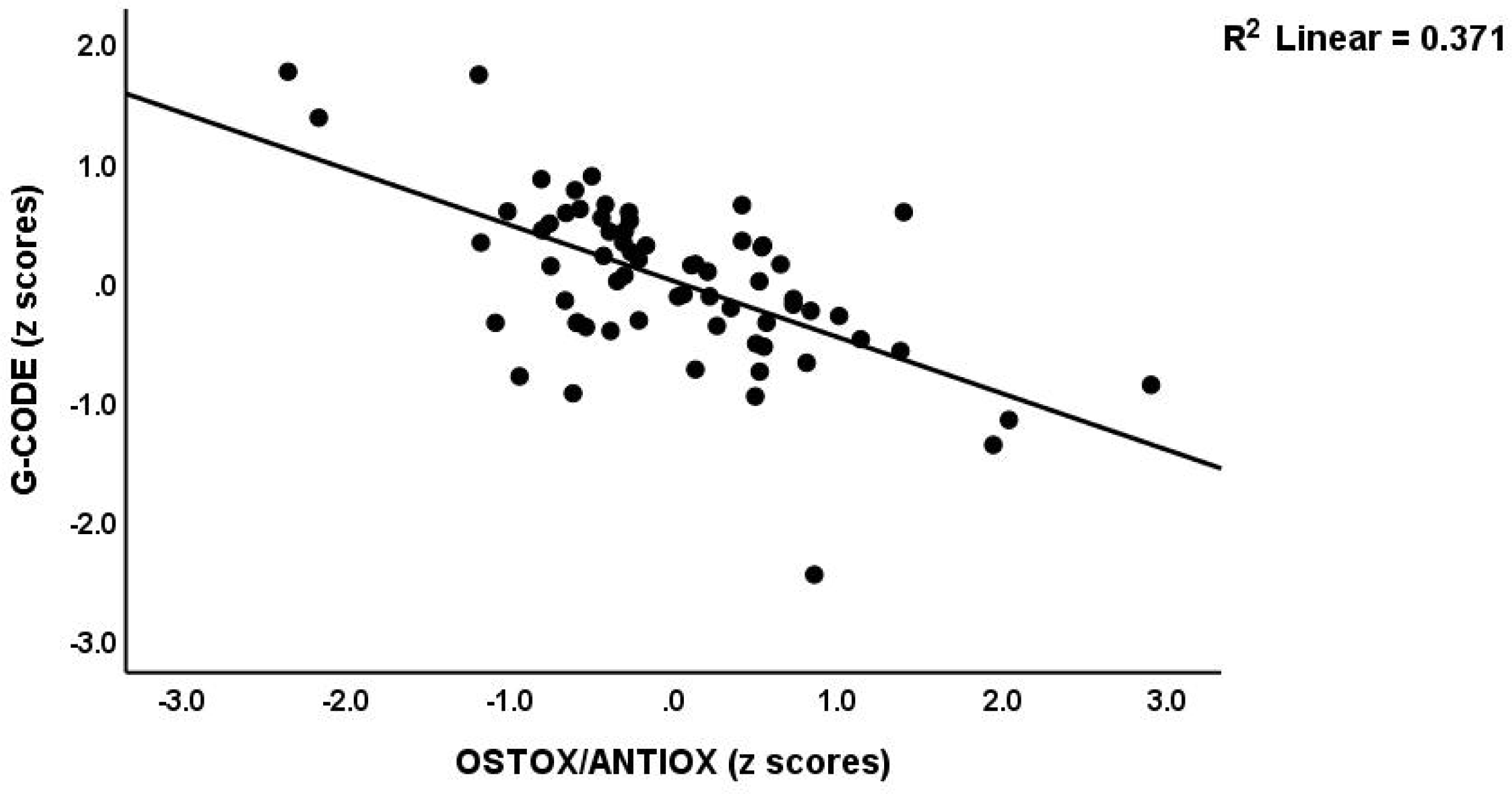
Partial regression of the general cognitive decline (G-CoDe) score on the oxidative stress toxicity / antioxidant (OSTOX/ANTIOX) ratio.

**Figure 2.**
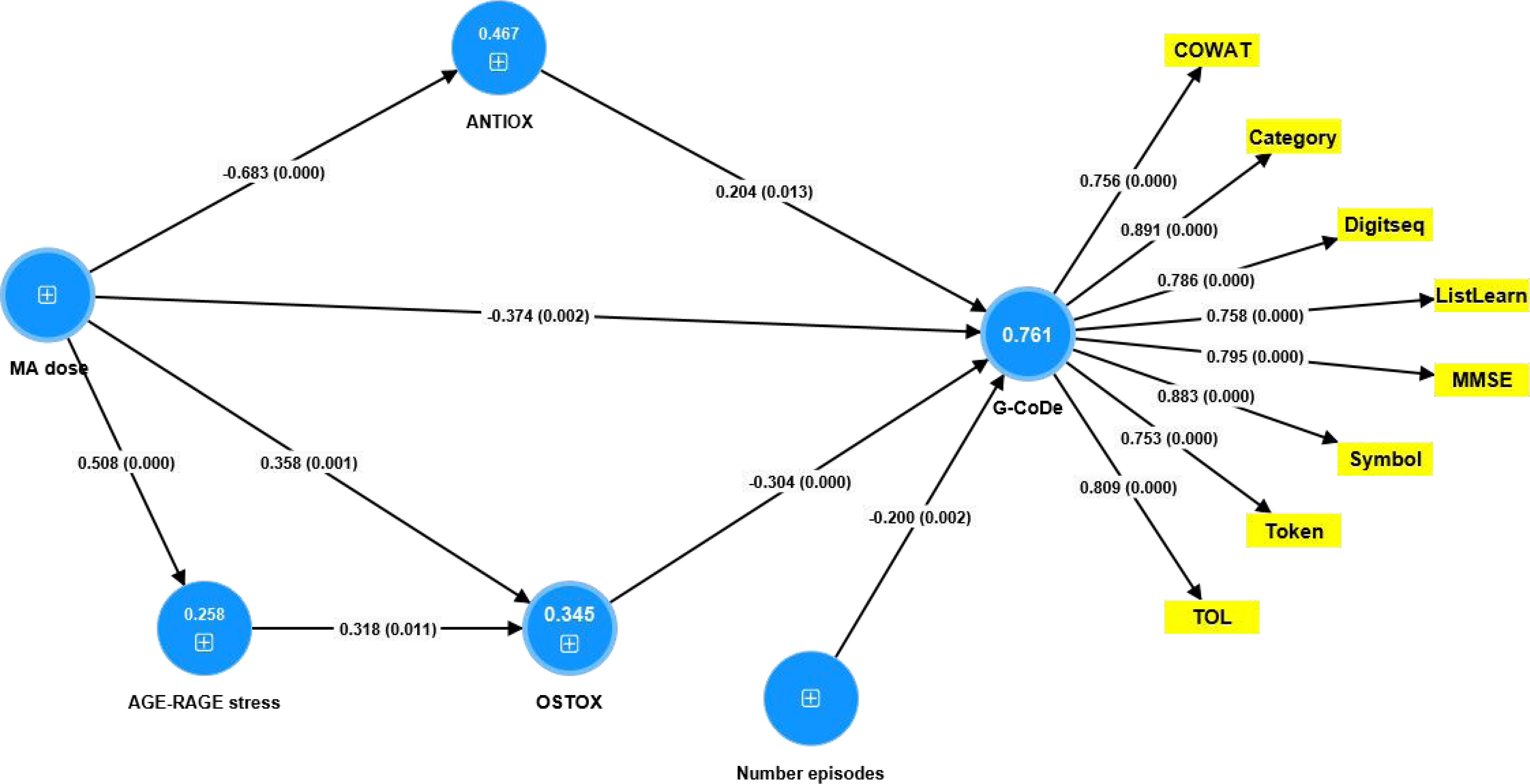
Results of partial least squares (PLS) analysis. The outcome variable is a factor extracted from 8 cognitive test scores. This factor is predicted by the number of psychotic episodes, methamphetamine (MA) use and dose, and two biomarkers, namely oxidative stress toxicity (OSTOX) and antioxidant defenses (ANTIOX). AGE-RAGE stress is allowed to predict OSTOX, whereas MA use and dose is allowed to predict all other variables. As such, the biomarkers mediate the effects of MA use and dose on the G-CoDe. AGE-RAGE: advanced glycation end products (AGEs) and AGE receptors; COWAT: Controlled Oral Word Association Test; Category: Test of Category instances; DigitSeq: Digit Sequencing Task; ListLearnL: List Learning test; MMSE: Mini Mental State Examination; Symbol: symbol coding; Token: Token Motor Task; TOL: Tower of London.

**Table 5.** Results of multiple regression analyses with the general cognitive decline (G-CoDe) scores as dependent variable and biomarkers as explanatory variables. G-CoDe: general cognitive decline (See Table 1), FTD+G-CoDE: factor based on formal thought disorders and the G-CoDe test scores; FPE+G-CoDe: factor based on formal thought disorders, psychosis, excitation and the G-CoDe test scores (see Table 2); OSTOX: oxidative stress toxicity; ANTIOX: antioxidant defenses; sRAGE: soluble receptors for advanced glycation end products.

| Dependent Variables | Explanatory Variables | Coefficients of input variables |  |  | Model statistics |  |  |  |
| --- | --- | --- | --- | --- | --- | --- | --- | --- |
| | | $\beta$ | t | p | R<sup>2</sup> | F | df | p |
| #1a. G-CoDe | **Model** |  |  |  | 0.647 | 38.44 | 3/63 | <0.001 |
|  | OSTOX/ANTIOX | -0.509 | -5.72 | <0.001 |  |  |  |  |
|  | Magnesium | 0.259 | 2.98 | 0.004 |  |  |  |  |
|  | sRAGE | -0.234 | -2.81 | 0.007 |  |  |  |  |
| #1b. G-CoDe | **Model** |  |  |  | 0.702 | 49.51 | 3/63 | <0.001 |
|  | OSTOX/ANTIOX | -0.486 | -6.09 | <0.001 |  |  |  |  |
|  | Number psychotic episodes. | -0.371 | -4.72 | <0.001 |  |  |  |  |
|  | sRAGE | -0.197 | -2.56 | 0.013 |  |  |  |  |
| #2. FTD+GCoDe | **Model** |  |  |  | 0.662 | 41.05 | 3/63 | <0.001 |
|  | OSTOX/ANTIOX | 0.501 | -5.54 | <0.001 |  |  |  |  |
|  | Magnesium | 0.265 | 3.10 | 0.003 |  |  |  |  |
|  | sRAGE | -0.253 | -3.10 | 0.003 |  |  |  |  |
| #3. FPE+GCoDe | **Model** |  |  |  | 0.658 | 40.67 | 3/63 | <0.001 |
|  | OSTOX/ANTIOX | 0.389 | 4.15 | <0.001 |  |  |  |  |
|  | Magnesium |  |  |  |  |  |  |  |
|  | sRAGE |  |  |  |  |  |  |  |

**Table 6.** Results of multiple regression analyses with the general cognitive decline (G-CoDe) score as dependent variable and biomarkers as explanatory variables. G-CoDe: general cognitive decline (See Table 1), FTD+G-CoDE: factor based on formal thought disorders and the G-CoDe tests; FPE+G-CoDe: factor based on formal thought disorders, psychosis, excitation, and the G-CoDe tests (see Table 2), MPO: myeloperoxidase.

| Dependent Variables | Explanatory Variables | Coefficients of input variables |  |  | Model statistics |  |  |  |
| --- | --- | --- | --- | --- | --- | --- | --- | --- |
| | | $\beta$ | T | p | R<sup>2</sup> | F | df | P |
| #1. G-CoDe | Model |  |  |  | 0.146 | 6.50 | 1/38 | 0.015 |
|  | MPO | -0.382 | -2.55 | 0.015 |  |  |  |  |
| #2. FTD+GCoDe | Model |  |  |  | 0.152 | 6.79 | 1/38 | 0.013 |
|  | MPO | -0.389 | -2.61 | 0.013 |  |  |  |  |
| #3. PEF+GCoDe | Model |  |  |  | 0.191 | 9.17 | 2/37 | 0.001 |
|  | Alcohol use | -0.413 | -3.06 | 0.004 |  |  |  |  |
|  | MPO | -0.376 | -2.79 | 0.008 |  |  |  |  |

### Results of PLS analysis

The PLS model with the G-Code as the final outcome variable is depicted in **Figure 1**. The latter was conceptualized as an extracted latent vector from the outcomes of eight neurocognitive tests. This factor, called G-CoDe, demonstrated satisfactory model fit data, as evidenced by Cronbach’s alpha = 0.922%, AVE = 0.649%, and composite reliability = 0.936. Every indicator variable was permitted to predict the G-CoDe. In addition, the number of episodes and MA dose were permitted to predict the biomarkers and the G-CoDe. As a result, a multistep mediation model was established. At 0.050, the model quality fit SRMR was sufficient. All Q2 predicted values for the manifest and latent variables were greater than zero, according to PLSPredict, and CVPAT analysis demonstrated that the model and every indicator exhibited superior performance than the naive benchmark. We established that 76.1 percent of the variance in the G-CoDe was accounted for by OSTOX, ANTIOX, number of episodes, and MA dose, according to bootstrapping. MA dose accounted for 46.7% of the variance in ANTIOX and AGE-RAGE stress explained 34.5% of the variance in OSTOX. Moreover, MA dose accounted for 25.8% of the variance in AGE-RAGE stress. A noteworthy specific indirect (mediated) effect of MA dose on G-CoDe was observed, with OSTOX (t=-2.74, p=0.006) and ANTIOX (t=2.14, p=0.032) as partial mediators. Additionally, OSTOX mediated a significant indirect effect of AGE-RAGE stress on G-Code (t=-2.07, p=0.038). MA dose (t=-9.89, p<0.001), OSTOX (t=-3.57, p<0.001), number of episodes (t=-3.05, p=0.001), ANTIOX (t=2.49, p=0.013), and AGE-RAGE stress (t=-1.97, p=0.049) all had significant total effects (in descending order of importance).

## Discussion

### Impaired neurocognitive tests and G-CoDe in MA use

The first major finding of this research is that a validated factor was identified from the seven BACS tests and the MMSE score. Furthermore, the factor score of this identified factor was significantly lower in patients with MA use as compared to the control group. In fact, patients with MA had lower scores on all eight cognitive test scores compared to the control group. Additionally, individuals who had experienced transient psychotic symptoms performed worse on the COWAT, symbol coding, and TOL tests than those who had not experienced psychotic symptoms. This latent construct calculated from the eight cognitive tests signifies a general cognitive decline (G-CoDe) among patients who have used MA for at least three months. Consequently, this G-CoDe is responsible for the manifestations of impairments observed in working memory (Digit Sequencing Task), verbal episodic memory (List Learning test), semantic fluency (Controlled oral Word Association test), attention (Symbol Coding), and executive functions (Tower of London).

The decreased levels observed in the neuropsychological test scores and the G-CoDe corroborate prior research indicating that neurocognitive assessments are compromised in MA users [15,21,22,32]. Furthermore, inhibitory control and decision-making are both impacted by MA dependence [33]. Bernheim et al. [15] established a correlation between the cognitive impairments that ensued after MA abuse and both the duration of drug use and the degree of dependence. In our study, however, no correlations were observed between the G-CoDe and any of the abuse characteristics (e.g., MA dose, number of psychoses, or SDS score) among the selected group of MA patients. However, following the introduction of biomarkers, G-CoDe was affected by MA use and dose.

According to research conducted by Hart et al. [21], the diminished cognitive test scores observed in MA abusers are still within the normal range. We determined in our research that the average MMSE score for patients with MA was approximately 25. In most cases, cognitive impairment is indicated by an MMSE score below 23 [34], whereas scores between 24 and 26 do not necessarily indicate significant impairment [35]. However, the MMSE scores for mild cognitive impairment (MCI) and deficit schizophrenia are 26.2 ±2.3 and 24.9 ±4.1, respectively [36]. The fact that subjects with MCI suffer from objective memory deficits suggests that such a minor deterioration in the MMSE could potentially have clinical ramifications. Moreover, in schizophrenia, decreased MMSE ratings are associated with increased ratings of psychotic symptoms, suggesting that these effects are clinically significant [25]. Consequently, there is clinical significance to the finding that MA users have lower MMSE and G-CoDe levels.

Prior observations showed that a G-CoDe could be extracted from neurocognitive test results in patients with schizophrenia [23,24,25,37]. For example, it was possible to develop a G-Code for schizophrenia by extracting a single factor from the seven BACS test scores [23,38,39]. Maes and Kanchanatwan et al. [25] extracted from executive factor scores (as measured by the Cambridge Automated Battery or CANTAB tests), the verbal fluency test (VFT), world list memory (WLM), True Recall, and the MMSE one validated and unidimensional factor. Hence, the G-CoDe observed in individuals with schizophrenia is like the G-CoDe determined in subjects who abuse MA.

### Associations between neurocognition and psychopathology

We found that patients exhibiting severe psychosis and excitation demonstrated substantially lower scores on the G-Code and three BACS tests, namely attention, executive functions, and verbal fluency, in comparison to those who did not exhibit psychosis and excitation. Furthermore, like the case of schizophrenia [25], correlations between the G-CoDe, FTD, psychosis, and excitation were found to be highly significant in the current study. According to Maes and Kanchanatawan [25], G-CODE accounted for 44.8% of the variability observed in the psychotic symptoms and FTD of schizophrenia. It was discovered in the present study that symptoms such as FTD, psychosis, and excitation belonged to the same G-Code factor constructed using neurocognitive test scores. Likewise, in schizophrenia, impairment is the MMSE, VFT, WLM, and executive functions belonged to the same factor extracted from psychosis, hostility, excitation, mannerism (PHEM), FTD, and psychomotor retardation [40].

However, it is crucial to emphasize that our assessments of the G-CoDe manifestations occurred after the resolution of the transient psychotic symptoms. Because MA abuse may cause a cognitive decline of unknown duration [32], it could be hypothesized that the G-CoDe is a vulnerability factor that may increase the propensity to develop psychosis during acute intoxication. However, to determine how long the G-CoDe may last and whether the lowered G-CoDe predicts psychotic episodes, future research should reexamine the G-CoDe manifestations several weeks to months after the transient psychotic episode.

In conclusion, the presence of correlations between the G-CoDe and psychotic symptoms, and FTD, seems to be a defining characteristic shared by MA-induced psychosis and schizophrenia. This finding could potentially support the notion that MA-induced psychosis serves as a model for schizophrenia. A significant distinction, however, is that psychosis induced by MA is most often a brief, transient condition, whereas schizophrenia is a chronic disorder.

### Impact of biomarkers on the G-CoDe in MA use

The second major finding of this study is that O&NS and AGE-RAGE stress and lowered antioxidant defenses are strongly associated with a decrease in the G-CoDe. Thus, 64.7% of the variance in the G-CoDe was accounted for by the increased O&NS and sRAGE, and decreased antioxidant levels, according to this study. Additionally, certain parallels can be drawn with schizophrenia in this context. Maes [24] conducted a review of various O&NS and antioxidant biomarkers that have been linked to the G-CoDe in schizophrenia. These biomarkers include elevated levels of LOOH and AOPP, as well as decreased antioxidant defenses, which include - SH groups, IgM responses to oxidative specific epitopes, and decreased paraoxonase 1 activity [40]. Thus, once more, certain parallels can be observed between the neurocognitive impairments associated with schizophrenia and those caused by MA.

Furthermore, the current investigation revealed, via PLS mediation analysis, that the impacts of MA dosage and usage on the G-CoDe were mediated in part by elevated levels of AGE-RAGE and O&NS stress and decreased antioxidant defenses. This suggests that the use of MA and its dosage induces detrimental outcome pathways (AGE-RAGE stress and O&N) and suppresses protective outcome pathways (antioxidant depletion), which ultimately may result in cognitive function abnormalities. Moreover, this mode of mediation resembles findings in schizophrenia. Thus, according to a study by Maes et al. [40], the main symptoms of schizophrenia and a factor composed of the G-CoDe are influenced by the PON1 gene in a manner mediated by increased oxidative stress and decreased antioxidant defenses.

There has been a hypothesis that the neurocognitive deficits in schizophrenia are underpinned by the same pathways that determine the psychotic symptoms [25]. Neurocognitive disorders of this nature are indicative of dysfunctions in “prefronto-temporal, prefronto-parietal, prefronto-striato-thalamic, hippocampal, and amygdal neural circuits,” as previously mentioned [25,41,42]. Cognitive impairments can be considered an immediate outcome of “compromised cerebral function” that occurs prior to the onset of psychotic symptoms [43,44]. Therefore, there exists a potential causal relationship between the PHEMN symptoms, FTD, and PMR of schizophrenia and these neurocognitive impairments [25]. The findings of the current study indicate that the G-CoDe is influenced by the neuro-oxidative pathways, which exert their toxic influence via various mechanisms. These mechanisms include disruptions in astroglial and neuronal projections, neurogenesis, neuroplasticity, synapse functions, protein regulation processes, cell signaling, apoptosis, and transcription [24,25]. Furthermore, AGEs are detrimental because they generate reactive oxygen species (ROS) and nitrogen species (RNS), in addition to inducing oxidative stress and inflammation [17]. These effects result in alterations to the structure and function of proteins, dysfunction, and apoptosis of cells, and ultimately damage to tissues (5). RAGE-mediated intracellular signaling can result in an upregulation of ROS and inflammatory processes upon ligand binding [45-48].

## Data Availability

The dataset generated and/or analyzed during the current research period will be provided upon reasonable request from the corresponding author (MM) and after the author fully utilizes the dataset.

## Acknowledgment

We thank the staff of the Psychiatry Unit of Al-Hussein Medical City in the Kerbala Governorate for their help in the collection of samples. We also acknowledge the work of the highly skilled staff of Asia Clinical *L*aboratory in Najaf for their help in the ELISA and spectroscopic measurements.

## Author’s contributions

All the contributing authors have participated in the preparation of the manuscript.

## Data Access Statement

The dataset generated during and/or analyzed during the current study will be available from the corresponding author (MM) upon reasonable request and once the dataset has been fully exploited by the authors.

## Ethical statement

Before participating in this study, all participants, and first-degree relatives of participants with MA SUD (legal representatives are the mother, father, brother, spouse, or son) gave written informed consent. The study was approved by the *institutional review board* (IRB) of the College of Science, University of Kufa, Iraq (89/2022), Karbala Health Directorate-Training and Human Development Center (Document No.18378/ 2021).

